# Beyond Accuracy: Investigating the Potential Clinical Utility of Predicting Functional Dependency and Severe Disability or Death in Successfully Reperfused Patients using Machine Learning

**DOI:** 10.1101/2020.11.17.20232280

**Authors:** Raphael Meier, Meret Burri, Samuel Fischer, Richard McKinley, Simon Jung, Thomas Meinel, Urs Fischer, Eike I. Piechowiak, Pasquale Mordasini, Jan Gralla, Roland Wiest, Johannes Kaesmacher

**Affiliations:** University Institute of Diagnostic and Interventional Neuroradiology, University Hospital Bern, Inselspital, University of Bern, Bern, Switzerland; Support Center for Advanced Neuroimaging, University Hospital Bern, Inselspital, University of Bern, Bern, Switzerland; Department of Neurology, University Hospital Bern, Inselspital, University of Bern, Bern, Switzerland; Department of Diagnostic, Paediatric and Interventional Radiology, University Hospital Bern, Inselspital, University of Bern, Bern, Switzerland

**Keywords:** Machine learning, modified Rankin Scale, functional impairment, clinical utility, acute ischemic stroke, deep learning

## Abstract

**Objectives:** Machine learning (ML) has been demonstrated to improve the prediction of functional outcome in patients with acute ischemic stroke. However, its value in a specific clinical use case has not been investigated. Aim of this study was to assess the clinical utility of ML models with respect to predicting functional impairment and severe disability or death considering its potential value as a decision-support tool in an acute stroke workflow.

**Materials and Methods:** Patients (n=1317) from a retrospective, non-randomized observational registry treated with Mechanical Thrombectomy (MT) were included. The final dataset of patients who underwent successful recanalization (TICI ≥ 2b) (n=932) was split in order to develop ML-based prediction models using data of (n=745, 80%) patients. Subsequently, the models were tested on the remaining patient data (n=187, 20%). For comparison, baseline algorithms using majority class prediction, SPAN-100 score, PRE score, and Stroke-TPI score were implemented. The ML methods included eight different algorithms (e.g. Support Vector Machines and Random forests), stacked ensemble method and tabular neural networks. Prediction of modified Rankin Scale (mRS) 3–6 (primary analysis) and mRS 5–6 (secondary analysis) at 3 months was performed using 25 baseline variables available at patient admission. ML models were assessed with respect to their ability for discrimination, calibration and clinical utility (decision curve analysis).

**Results:** Analyzed patients (n=932) showed a median age of 74.7 (IQR 62.7–82.4) years with (n=461, 49.5%) being female. ML methods performed better than clinical scores with stacked ensemble method providing the best overall performance including an F1-score of 0.75 ± 0.01, an ROC-AUC of 0.81 ± 0.00, AP score of 0.81 ± 0.01, MCC of 0.48 ± 0.02, and ECE of 0.06 ± 0.01 for prediction of mRS 3–6, and an F1-score of 0.57 ± 0.02, an ROC-AUC of 0.79 ± 0.01, AP score of 0.54 ± 0.02, MCC of 0.39 ± 0.03, and ECE of 0.19 ± 0.01 for prediction of mRS 5–6. Decision curve analyses suggested highest mean net benefit of 0.09 ± 0.02 at a-priori defined threshold (0.8) for the stacked ensemble method in primary analysis (mRS 3–6). Across all methods, higher mean net benefits were achieved for optimized probability thresholds but with considerably reduced certainty (threshold probabilities 0.24–0.47). For the secondary analysis (mRS 5–6), none of the ML models achieved a positive net benefit for the a-priori threshold probability 0.8.

**Conclusions:** The clinical utility of ML prediction models in a decision-support scenario aimed at yielding a high certainty for prediction of functional dependency (mRS 3–6) is marginal and not evident for the prediction of severe disability or death (mRS 5–6). Hence, using those models for patient exclusion cannot be recommended and future research should evaluate utility gains after incorporating more advanced imaging parameters.

## Introduction

The HERMES data and recent late-time window thrombectomy studies suggested a low number needed to treat with mechanical thrombectomy for improving the functional status of patients with an acute ischemic stroke due to a large vessel occlusion (NNT=2.6 in HERMES)^1^. The inclusion criteria of trials included in the HERMES meta-analysis were relatively strict. However, the large effect sizes observed, promising results derived from observational data in borderline indication groups^2–4^, and a relatively low procedural complication risk^1,5,6^, have been put forward as a major argument widening the indication criteria for mechanical thrombectomy (MT). This is also reflected by statements from the recent ESO/ESMINT guidelines for borderline indications group, suggesting that MT may be reasonable in patients with e.g. low ASPECTS or low NIHSS scores, if inclusion into randomized trials is not possible^7^.

Correspondingly, there was a considerable shift from carefully selecting patients to receive MT (rule in) towards treating the vast majority of patients, except for cases with convincing reasons to withhold MT (rule out). Such a paradigm shifts makes the treatment available for more patients^8–10^, but also inevitably comes with the risk of increasing futile interventions without a clinical benefit^11–13^. If one would be able to precisely determine which patient has a poor outcome despite a technically successful treatment, this can be used to assess the cost-benefit relationship of the treatment and enhance patient-oriented informed decision (e.g. withholding treatment in a patient with legally binding declaration of not wanting to live in moderate or severe dependency).

In the past, many different prediction approaches based on clinical variables at baseline have been introduced in patients with acute ischemic stroke, e.g. simple clinical scores such as the Pittsburgh Response to Endovascular (PRE) therapy score^14^, the Stroke-Thrombolytic Predictive Instrument (Stroke-TPI)^15^, and more recently prediction models based on Machine Learning^16,17^. Machine Learning (ML) methods showed potential to improve prediction performance when compared to clinical scores^18^. However, a recent systematic review^19^ has pointed out the following weaknesses of ML studies conducted so far: Small cohorts (median sample size is 475), no or only simple data imputation (e.g. median imputation), insufficient reporting of hyperparameter tuning, and a strong focus on discrimination with only three studies discussing calibration and no studies discussing clinical utility in context of a possible scenario-based integration into acute stroke care^20^.

Consequently, we set out to evaluate the performance of a wide variety of Machine Learning algorithms (including logistic regression, Support Vector Machines, Random forests) and tabular neural networks for predicting functional dependency and severe disability or death in patients with successful reperfusion. A particular aim of this study was the assessment of ML models with respect to their clinical utility (i.e. excluding patients from MT based on high likelihood for futile reperfusion) in pre-interventional prediction of 3-months functional impairment.

## Methods

### Study Design

We included patients (n=1317) of a single center from a retrospective, non-randomized observational registry purposed to investigate the safety and efficacy of second-generation market-released device for mechanical thrombectomy^1^. Details on the inclusion criteria of the registry were published previously^21^. We excluded patients with age below 18 years (n=4), patients with invalid values in clinical variables (n=2), patients who did not exhibit a proximal large vessel occlusion (n=121), patients with missing TICI score (n=1) and missing 3-months mRS (n=71). Furthermore, analysis was restricted to patients who were successfully recanalized (i.e. patients with TICI < 2b were excluded; n=186). Therefore, the developed models are purposed to predict 3-month functional impairment at baseline under the assumption of successful recanalization (TICI ≥ 2b). Figure 2 provides an overview on the intended use of the ML prediction in an acute stroke workflow. Approval by local ethics committee is available (Bernese/Swiss Stroke Registry: Kantonale Ethikkommission für die Forschung Bern, Bern, Switzerland, amendment access number: 231/2014 and BEYOND- SWIFT registry, access number: 2018-00766).

**Figure 1.**
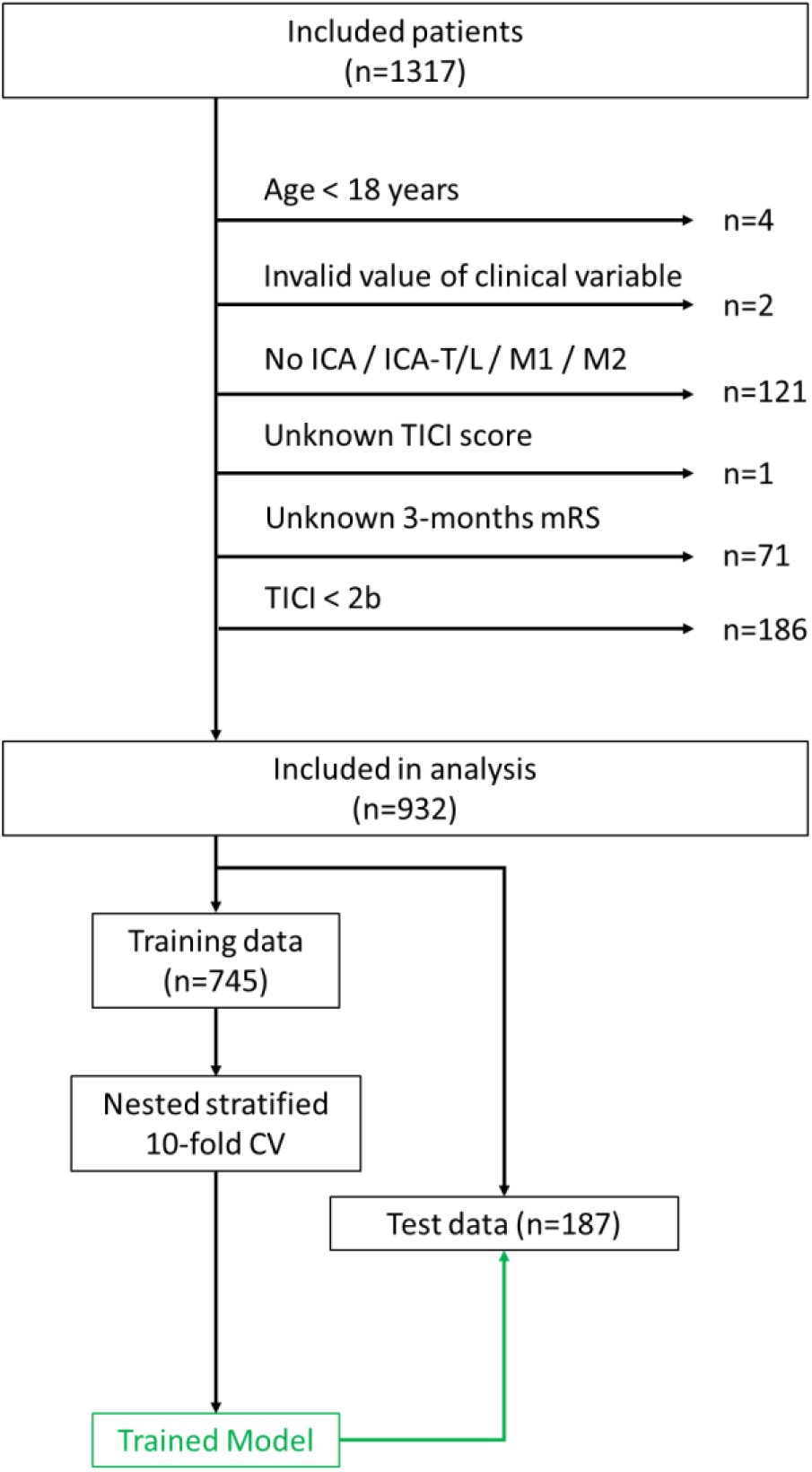
Flow chart of patient inclusion and experimental study design. Model development was based on nested stratified 10-fold cross-validation (CV) using training data (N=745). Trained models were evaluated on separate test set (n=187).

**Figure 2.**
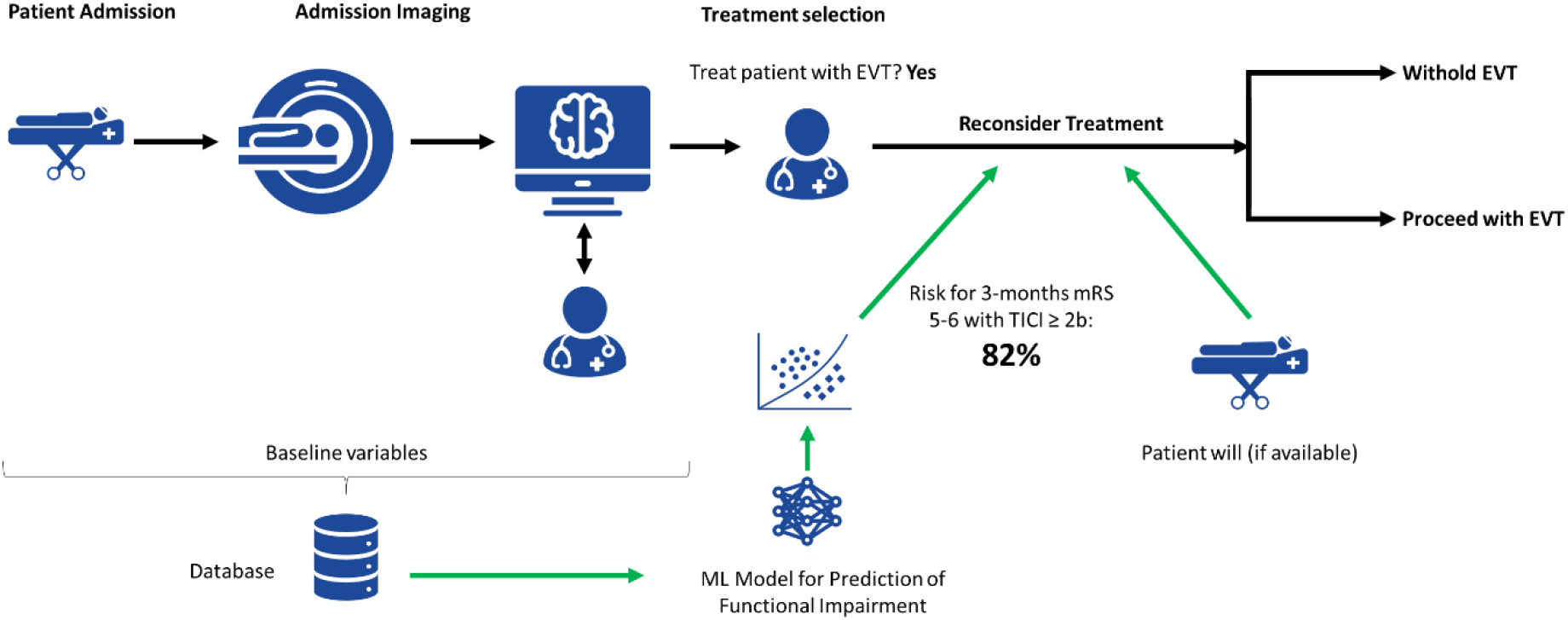
Intended use of ML prediction in a stroke workflow. The ML model outputs a probability (risk score) for mRS 5–6 based on variables available ahead of intervention. This information is provided to the treating physicians after selection of patient for EVT (indicated with green arrows) and could serve as a marker for futile recanalization.

The final dataset (n=932) was split randomly into a training set (80%, n=745) used for model development and an internal test set (20%, n=187). The flow chart describing patient inclusion and study design is shown in Figure 1.

### Baseline Variables

An initial set of 30 variables which are available on admission was included. Input variables (features) were dropped when more than 25% of the values were missing. Dropped features included risk due to coronary heart disease, systolic & diastolic blood pressure, admission international normalized ratio (INR), and admission platelet.

Therefore, we considered a total of 25 features available on admission including age on admission (continuous, years), sex (binary), direct vs transfer (binary), pre stroke independence (binary), risk due to diabetes (binary), risk due to arterial hypertension (binary), risk due to dyslipidemia (binary), risk due to smoking (binary), risk due to previous stroke (binary), medication anticoagulation (categorical), medication antiplatelet (categorical), medication statin (binary), admission glucose (continuous, mmol/L), quality symptom onset (categorical), wake up stroke (binary), in hospital stroke (binary), IVT bridging (binary), type of admission imaging (binary), site of occlusion (categorical), ASPECTS (DWI/CT, treated as continuous), tandem occlusion (binary), dissection (binary), NIHSS on admission (treated as continuous), time symptom onset to admission (continuous, min), time admission to groin puncture (continuous, min).

### Functional Outcome

Functional impairment was defined by the modified Rankin Scale (mRS) 3–6 at 3 months. The mRS was dichotomized to serve as binary target variable. The primary analysis included the prediction of mRS 3–6 at 3 months. Despite the fact that such a definition of futile reperfusion is highly debatable, the endpoint was chosen to make the results comparable to reported results from other studies. For the intended use case (Figure 2), a secondary analysis included prediction of mRS 5–6 at 3 months. This secondary endpoint more closely reflects true futile reperfusion, because quality of life may be substantial for patients with mRS grades 3 and 4^22^, while five-year quality-adjusted-life-expectancy in stroke survivors with mRS 5 is overall low (0.06)^22–24^.

Therefore, prediction of 3-months functional impairment based on clinical variables available at baseline was regarded as a binary classification problem (mRS ≤ 2 vs. mRS > 2 and mRS < 5 vs. mRS ≥ 5, respectively). The classes in the primary analysis (mRS ≤ 2 vs. mRS > 2) were roughly balanced (n=480, 51.5% mRS > 2), whereas classes in the secondary analysis (mRS < 5 vs. mRS ≥ 5) were imbalanced (n=245, 26.3% mRS ≥ 5).

### Machine Learning Methods

Missing values for input features were imputed using k-Nearest Neighbor (k-NN) imputer with k=15 neighbors on normalized feature data of the training and testing data separately. Categorical variables were one-hot encoded for all ML methods except the tabular neural network which relied on categorical embeddings. Feature values were normalized to [0,1] interval using min-max normalization.

Four baseline algorithms including Majority classifier, SPAN-100 score^25^ (age on admission + NIHSS on admission), PRE score^14^ (function of age on admission, NIHSS on admission and ASPECTS) and Stroke-TPI score^15^ were implemented. Stroke-TPI corresponds to a logistic regression on age on admission, NIHSS on admission, ASPECTS, and admission glucose.

Eight baseline ML algorithms including k-NN classifier using Manhattan distance, linear soft-margin SVM based on liblinear, linear soft-margin SVM based on libsvm, non-linear SVM with Radial Basis Function (RBF) kernel, regularized logistic regression with interaction terms, Gradient Boosting (XGBoost classifer), random forest (RF), and Multi-layer Perceptron (MLP) were implemented using Python’s (3.7.7) scikit learn^26^ (0.22.1), xgboost^27^ (1.2.0, for XGBoost classifier), and imbalanced-learn^28^ (0.6.2, for random forest) modules. Class imbalance was tackled using class weights for models in scikit learn and majority class undersampling of each bootstrap sample for the random forest. Calibration of the SVM models was improved using Platt’s scaling^29^.

The input features included both continuous and categorical (binary) variables. Therefore, we propose an ensemble method which includes a dedicated ML algorithm for each type of variable. In particular, a linear soft-margin SVM is used to process continuous variables only, a Boolean soft-margin SVM using a Tanimoto kernel function^30^ is employed to process the one-hot encoded categorical variables and a random forest is used to operate on all variables. The rationale is that by treating continuous variables and categorical variables separately, we obtain feature spaces with less distorted geometries and thus may potentially improve discrimination between classes for SVMs. The role of the RF is to capture non-linear relationships between features and target variable as well as interactions between input features. The three models (linear SVM, Boolean SVM, and RF) are combined in an ensemble by stacking their probabilistic outputs using a logistic regression model with l2-regularization (to prevent one model from dominating the final decision). Class imbalance was tackled using class weights on the level of the individual models and the stacking method.

Finally, we implemented a tabular neural network based on fast.ai’s tabular learner using categorical embeddings^31^. The architecture of the neural network is presented in the Supplementary Materials (Section Hyperparameter Optimization). The loss function was cross entropy with label smoothing to improve the calibration of the final model^32^. Class weights were used in the loss function computation to tackle class imbalance. The model was trained for 80 epochs using Ranger optimizer^33^ with a learning rate of 4e-03 (based on learning rate finder) and flat cosine annealing^34^. Early stopping^35^ based on the f1-score in the validation set was employed to prevent overfitting.

### Machine Learning Model Development & Testing

We used a nested, stratified 10-fold cross-validation strategy for model development. In the outer CV loop, the training dataset was split into 10 equally sized subsets. Nine out of the ten subsets were used for training and one for testing. In the inner loop, hyperparameter optimization was performed based on maximization of f1-score in a 10-fold randomized gridsearch using the data of the previously formed nine folds. A detailed overview on the hyperparameters of the ML algorithms is shown in the Supplementary Table 1.

For model testing, we trained ML algorithms using the setting of the nested CV’s inner loop on the complete training data (10-fold randomized grid search) and applied the resulting ML model to separate test data (n=187). We repeated this process 20 times using a different random seed for algorithm initialization in each run. It has been shown that performance of Deep Learning methods varies considerably depending on the choice of random seed^36^, and thus our intention was to capture this aspect.

### Statistical analysis

Univariate associations of clinical variables with functional outcome were assessed using Mann-Whitney U-test for continuous variables and Chi-square test for categorical variables (for primary analysis only). The statistical analysis of the different ML methods investigated model discrimination, calibration and clinical utility. Discrimination refers to the ability of the ML method to separate the two classes. Calibration refers to the ability of the ML model to provide accurate probability estimates. Finally, clinical utility can be regarded as a combination of discrimination and calibration with added clinical context in form of an a-priori defined threshold on predicted risk of functional impairment. In order to assess the ML models ability to discriminate at the threshold of p=0.5, we employed accuracy, balanced accuracy, precision, recall, f-1 score, specificity, and Matthew’s correlation coefficient (MCC). In addition, the Area Under the Curve (AUC) of the Receiver operating characteristic and the Average Precision (AP) score are reported as measures across all possible thresholds. The calibration of ML models was quantified using the Brier score and Expected Calibration Error (ECE) based on 10 bins. Feature importance was computed for all methods using the permutation method^37^ on the test set. We defined permutation feature importance through the decrease in f1-score when the values of a single feature are randomly shuffled. Decision curve analysis^38^ was employed to quantify the clinical utility of the ML models on test data. In particular, we defined a-priori a probability threshold of p=0.8 for risk of functional impairment (mRS 3–6) and severe disability or death (mRS 5–6) to assess net benefit (=difference between fraction of true positives and false positives, in which the latter are weighted by odds of probability threshold). In addition, we determined optimal probability thresholds for all models during model development to maximize f1-score. Net benefit for f1-optimized probability thresholds was reported as well. Results are reported as mean ± 1 standard deviation based on the stratified 10-fold cross-validation for model development. Performance on test data was reported as mean ± 1 standard deviation over 20 runs with different random seeds for algorithm initialization.

## Results

### Baseline characteristics

The median age of included patients (N=932) was 74.7 (62.7–82.4) years with (n=461, 49.5%) being female. Baseline characteristics of patients in training and test set are shown in Table 1. Univariate associations of clinical variables with functional outcome was assessed for both training and test data. For training data, age on admission, admission glucose, NIHSS on admission, ASPECTS, time symptom onset to admission, pre stroke independence, risk factor diabetes, risk factor arterial hypertension, risk factor smoking, anticoagulation medication, antiplatelet medication, IVT bridging, type of imaging, and site of occlusion were significantly different between patients with mRS ≤ 2 and patients with mRS > 2 (P < 0.01). For the test data, significant associations of ASPECTS, time symptom onset to admission, risk factor arterial hypertension, risk factor smoking, anticoagulation medication, antiplatelet medication, IVT bridging and site of occlusion were not present. However, time admission to groin puncture was significantly different between patients mRS ≤ 2 and patients with mRS > 2 (P < 0.01).

**Table 1.**
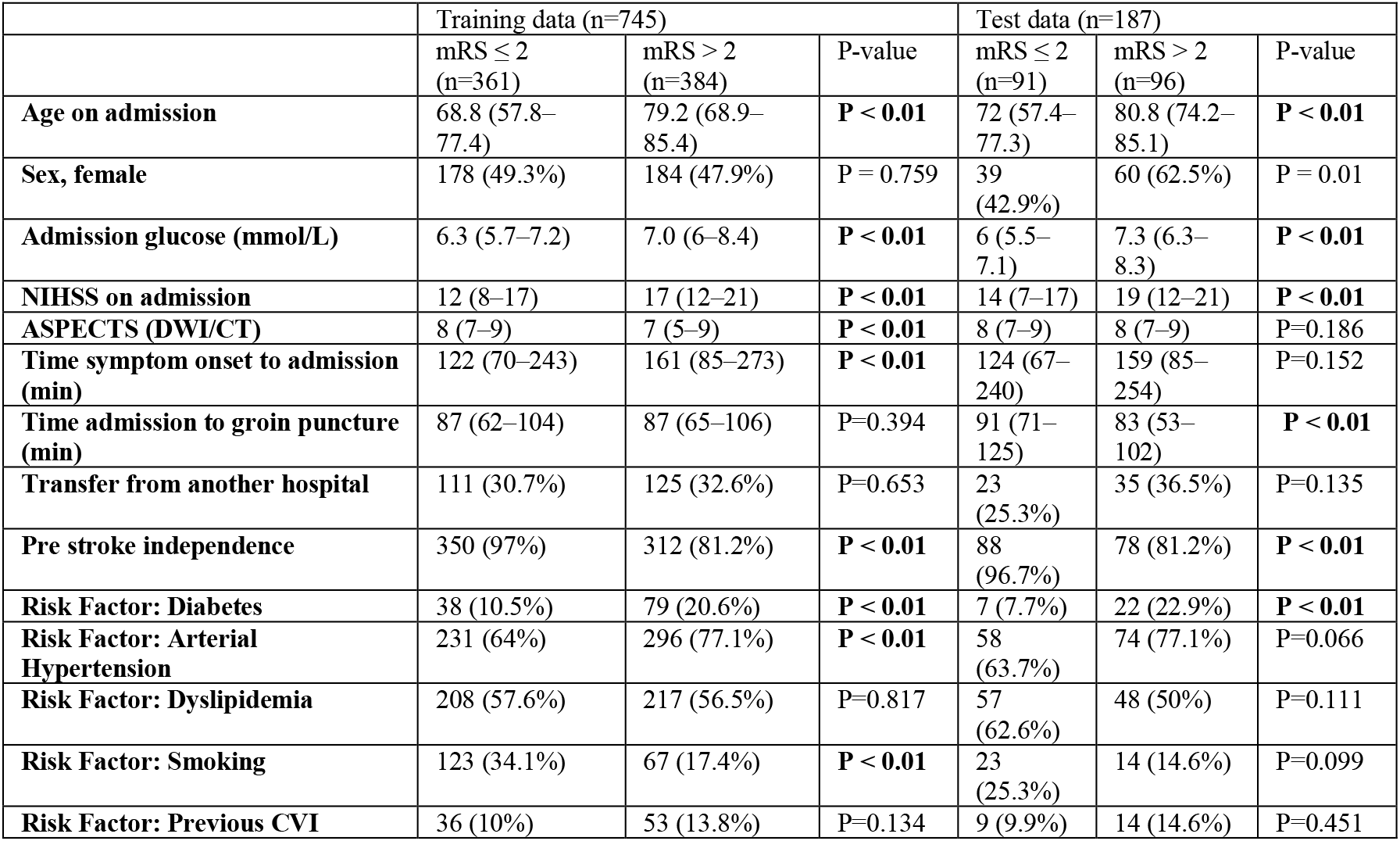
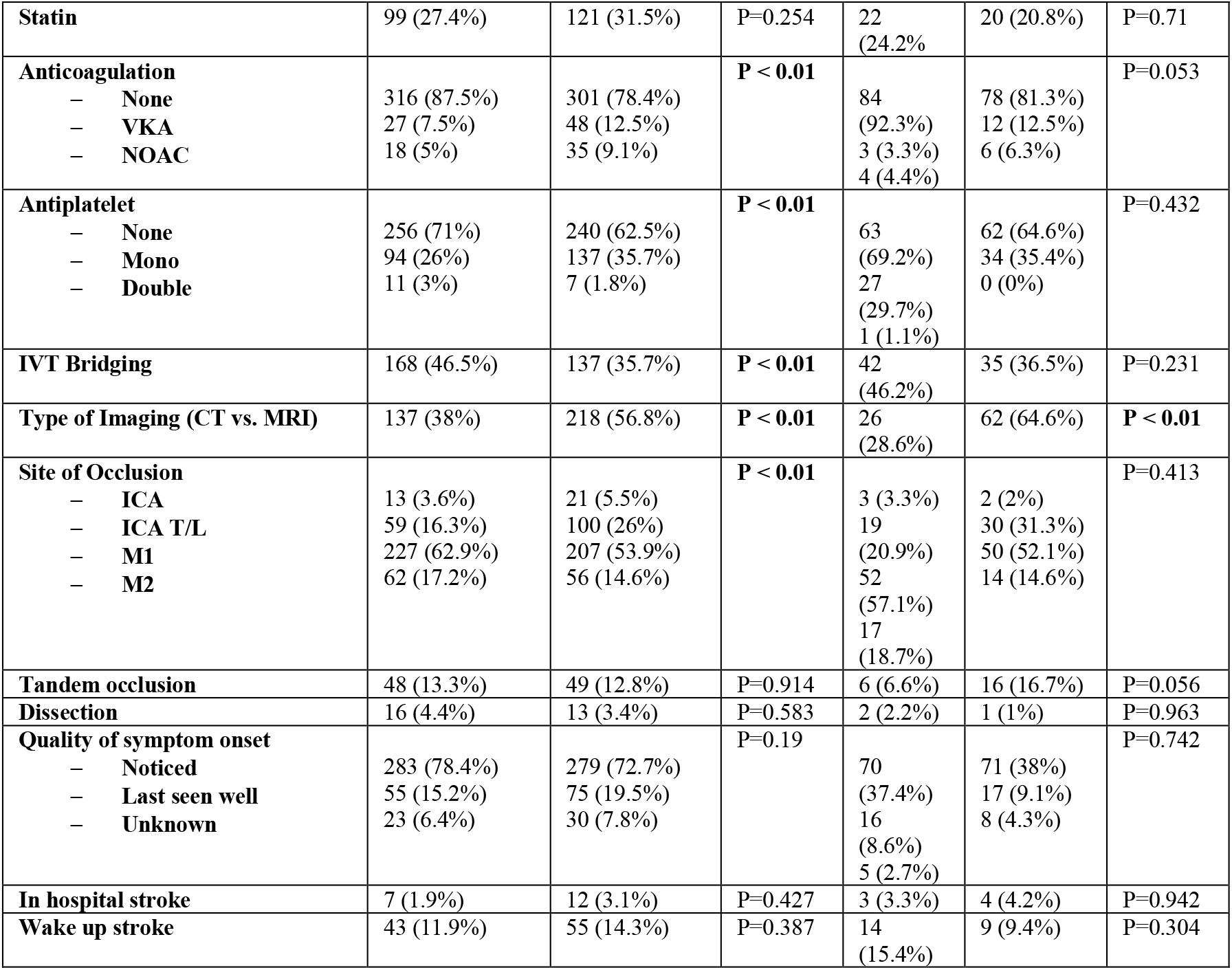
Baseline characteristics of all included patients (shown for primary analysis). Continuous variables were reported as median and interquartile range (IQR). Categorical variables were reported as counts (proportions).

### Prediction of Functional Impairment and Severe Disability or Death

Discrimination of ML methods (except for k-NN classifier) for prediction of mRS 3–6 at 3 months was superior when compared to naïve baseline algorithms (majority classifier, SPAN- 100 score, PRE score, Stroke-TPI score). Performance of all ML methods (except for k-NN classifier) was in a similar range. An overview of the results is provided in Table 2 (full data presented in Supplementary Table 2). Best overall performance on test data was achieved by the stacked ensemble method with F1-score of 0.75 ± 0.01, ROC-AUC of 0.81 ± 0.00, AP score of 0.81 ± 0.01, MCC of 0.48, and ECE of 0.06 ± 0.01. Computation of permutation importance revealed that among all methods (except k-NN classifier) the most important features included age on admission, NIHSS on admission, and pre stroke independence. The feature importance of the stacked ensemble method are shown in Figure 3 (see Supplementary Figures 1 & 2 for other methods and for secondary analysis).

**Table 2.**
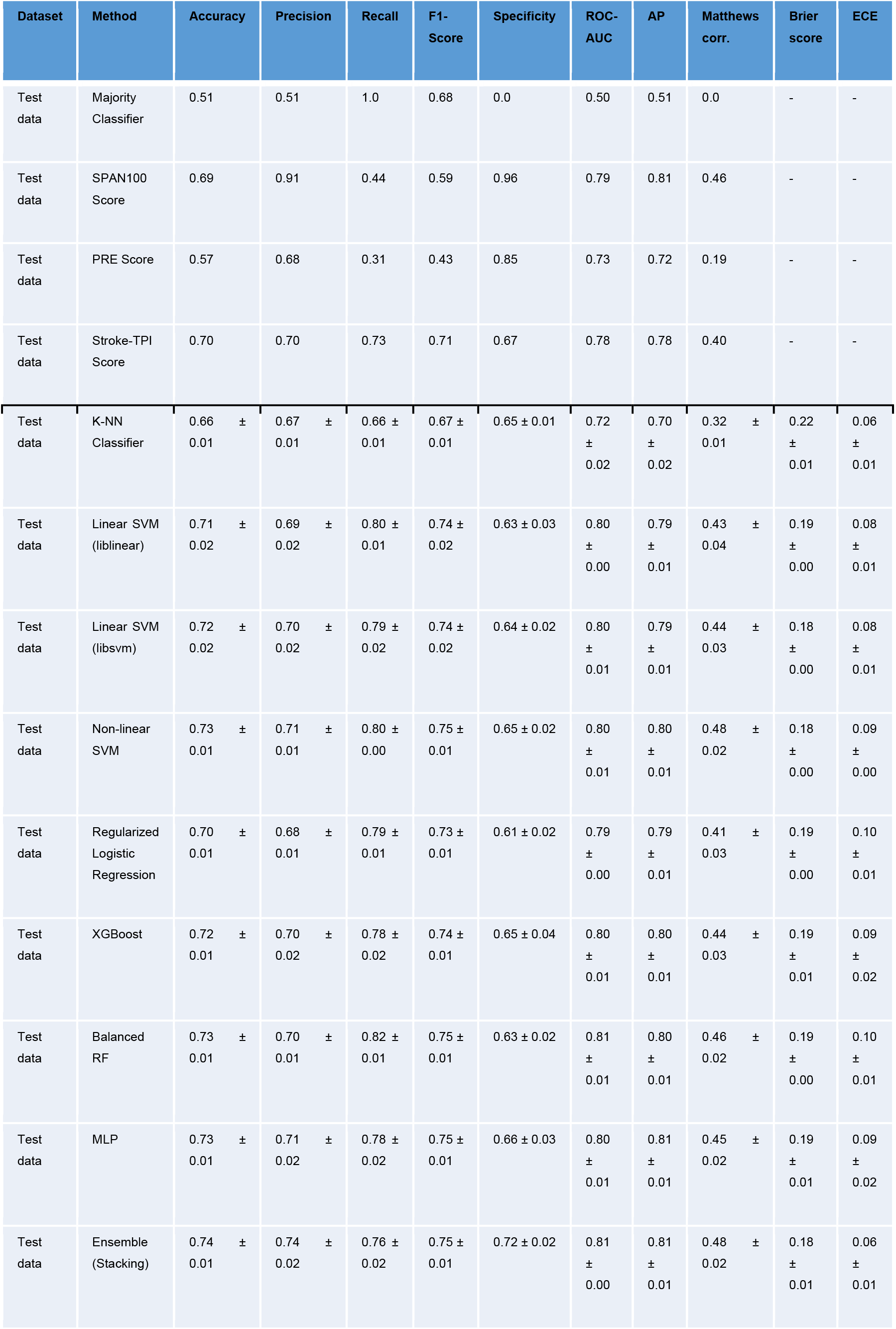
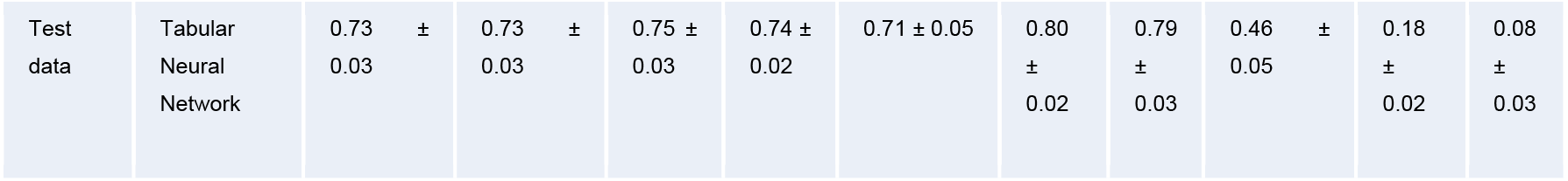
Performance for mRS 3–6 prediction. Values are reported as mean ± std. Brier score and ECE are reported for ML algorithms only. (for results of 10-fold cross-validation on training data, see Supplementary Table 2)

**Figure 3.**
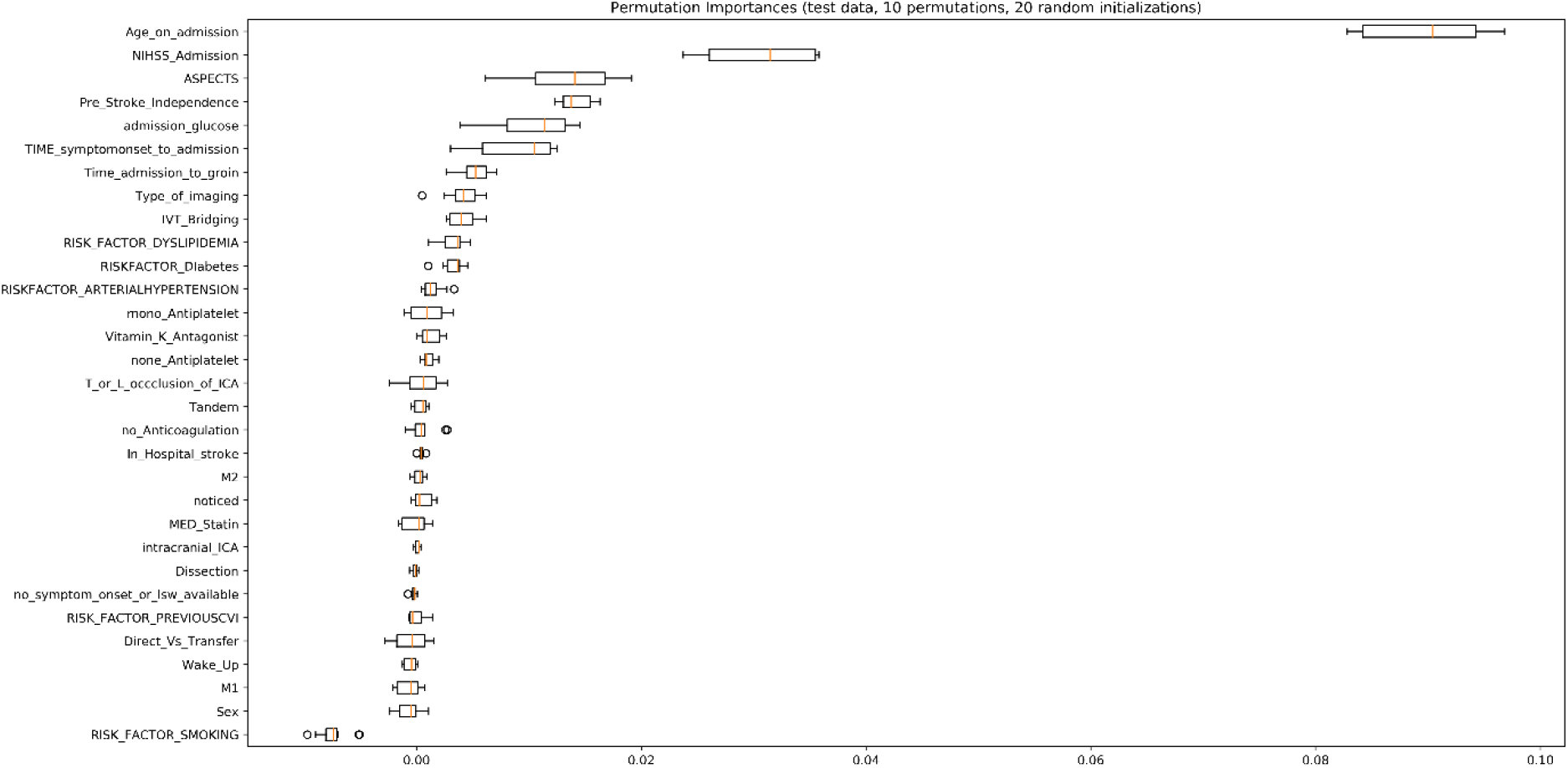
Permutation feature importance computed for the stacked ensemble method (mean values of 10 feature permutations and 20 random initializations).

When looking at the performance for prediction of mRS 5–6 at 3 months (secondary analysis) a considerable drop in performance for all ML methods can be observed. The stacked ensemble method achieved F1-score of 0.57 ± 0.02, ROC-AUC of 0.79 ± 0.01, AP score of 0.54 ± 0.02, MCC of 0.39 ± 0.03, and ECE of 0.19 ± 0.01 on the test set. An overview of the results for the secondary analysis can be found in the Supplementary Table 3.

### Clinical Utility of Machine Learning Models

Decision curve analysis was performed for Stroke-TPI score and all ML methods. It included the mean net benefit for optimized probability thresholds (maximum F1-score) and mean net benefit for a-priori defined probability threshold of p=0.8. The results of the primary analysis (mRS 3–6) are shown in Table 3. Exemplary decision curves for regularized logistic regression, stacked ensemble method, and tabular neural network are shown in Figures 4–6 (decision curves of all methods are displayed in the Supplementary Figure 3). Highest mean net benefit of 0.09 ± 0.02 for p=0.8 was achieved by the stacked ensemble method. Optimized probability thresholds provided higher mean net benefits across all methods but with considerably reduced probability thresholds (p=0.24–0.47). For the secondary analysis (mRS 5–6), none of the ML models achieved a positive net benefit for the threshold p=0.8 (see Supplementary Table 2). Similarly to the primary analysis, the optimized thresholds provided improved mean net benefits for lower probability threshold values (p=0.22–0.57). Detailed results of secondary analysis are provided in the Supplementary Materials (Supplementary Table 4 and Supplementary Figure 4).

**Table 3.** Results of decision curve analyses for different ML algorithms (and Stroke-TPI score) in case of mRS 3– 6 prediction reported for the f1-optimized probability thresholds and the a-priori defined clinically relevant threshold of p=0.8.

| Method | Mean Net Benefit for F1-optimized threshold | Mean Net Benefit for $p=0.8$ |
| --- | --- | --- |
| Stroke-TPI score | $0.28 \pm 0.00$ for $p=0.40$ | $0.04 \pm 0.00$ |
| k-NN Classifier | $0.36 \pm 0.00$ for $p=0.24$ | $-0.02 \pm 0.03$ |
| Linear SVM (liblinear) | $0.26 \pm 0.01$ for $p=0.46$ | $0.00 \pm 0.02$ |
| Linear SVM (libsvm) | $0.27 \pm 0.01$ for $p=0.45$ | $0.05 \pm 0.01$ |
| Non-linear SVM | $0.29 \pm 0.00$ for $p=0.38$ | $0.03 \pm 0.02$ |
| Regularized Logistic Regression | $0.26 \pm 0.01$ for $p=0.45$ | $0.01 \pm 0.02$ |
| XGBoost | $0.30 \pm 0.01$ for $p=0.40$ | $0.06 \pm 0.03$ |
| Balanced Random Forest | $0.26 \pm 0.01$ for $p=0.47$ | $0.06 \pm 0.02$ |
| MLP | $0.31 \pm 0.01$ for $p=0.37$ | $0.01 \pm 0.05$ |
| Ensemble (Stacking) | $0.30 \pm 0.01$ for $p=0.4$ | **$0.09 \pm 0.02$** |
| Tabular Neural Network | $0.27 \pm 0.04$ for $p=0.44$ | $0.06 \pm 0.04$ |

**Figure 4.**
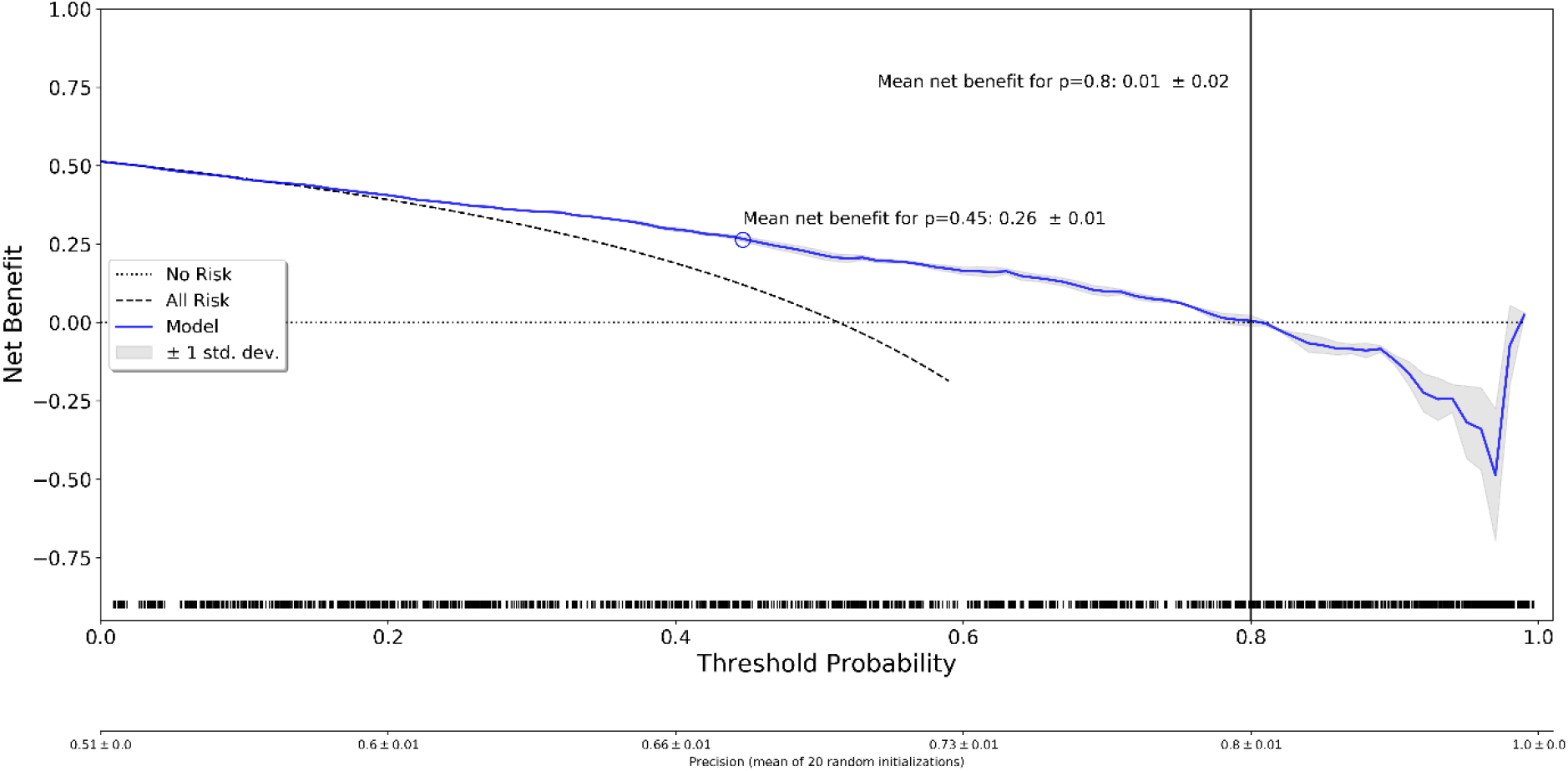
Decision curve analysis for regularized logistic regression. The blue line indicates the mean and the gray shaded region indicates ± 1 std. band over 20 random initializations. The rug plot shows the samples used for computation of the decision curve. ‘No risk’ denotes the trivial strategy of always predicting mRS 0–2, and ‘All Risk’ denotes the strategy of always predicting mRS 3–6.

**Figure 5.**
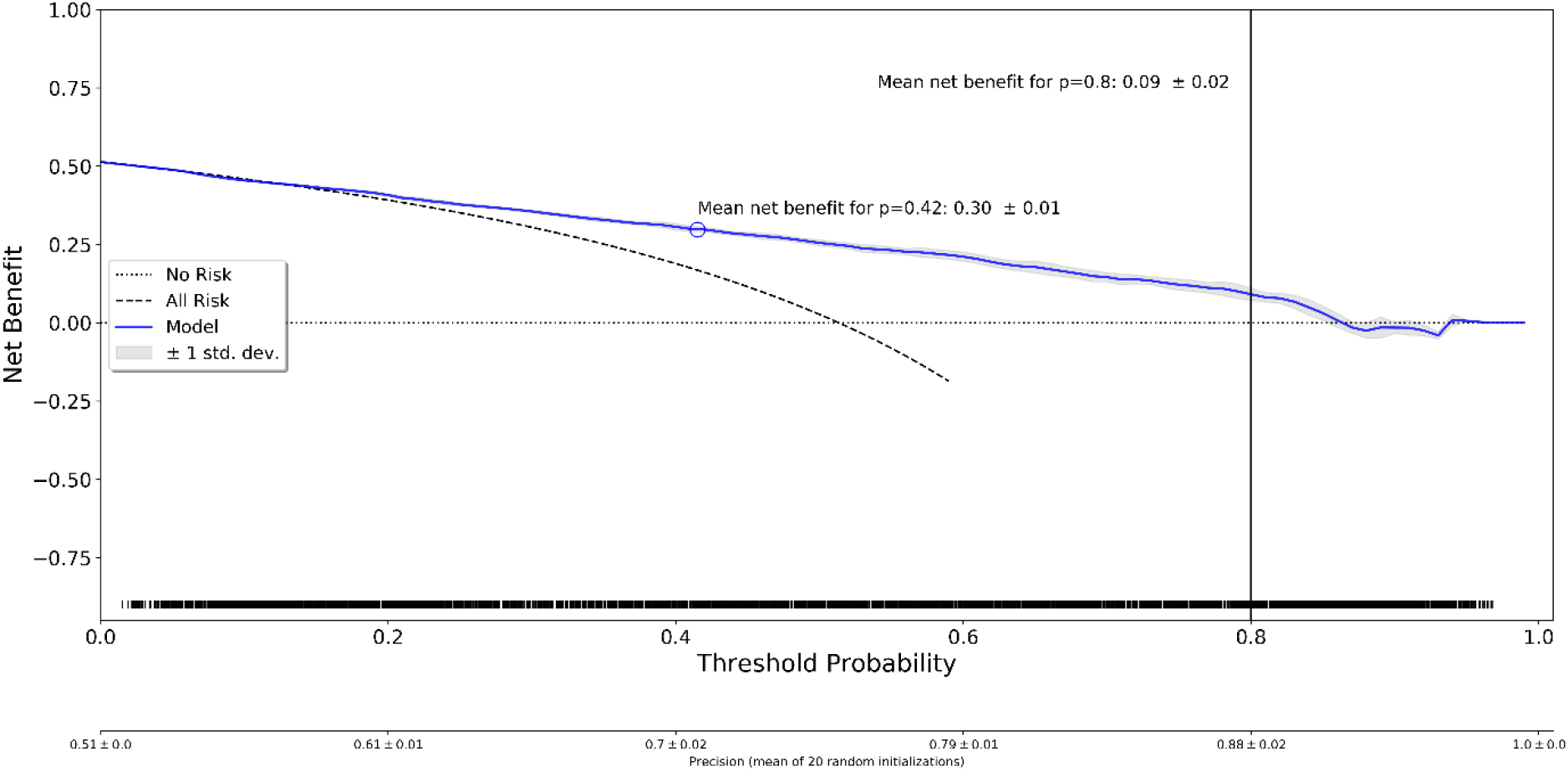
Decision curve analysis for stacked ensemble method. The blue line indicates the mean and the gray shaded region indicates ± 1 std. band over 20 random initializations. The rug plot shows the samples used for computation of the decision curve. ‘No risk’ denotes the trivial strategy of always predicting mRS 0–2, and ‘All Risk’ denotes the strategy of always predicting mRS 3–6.

**Figure 6.**
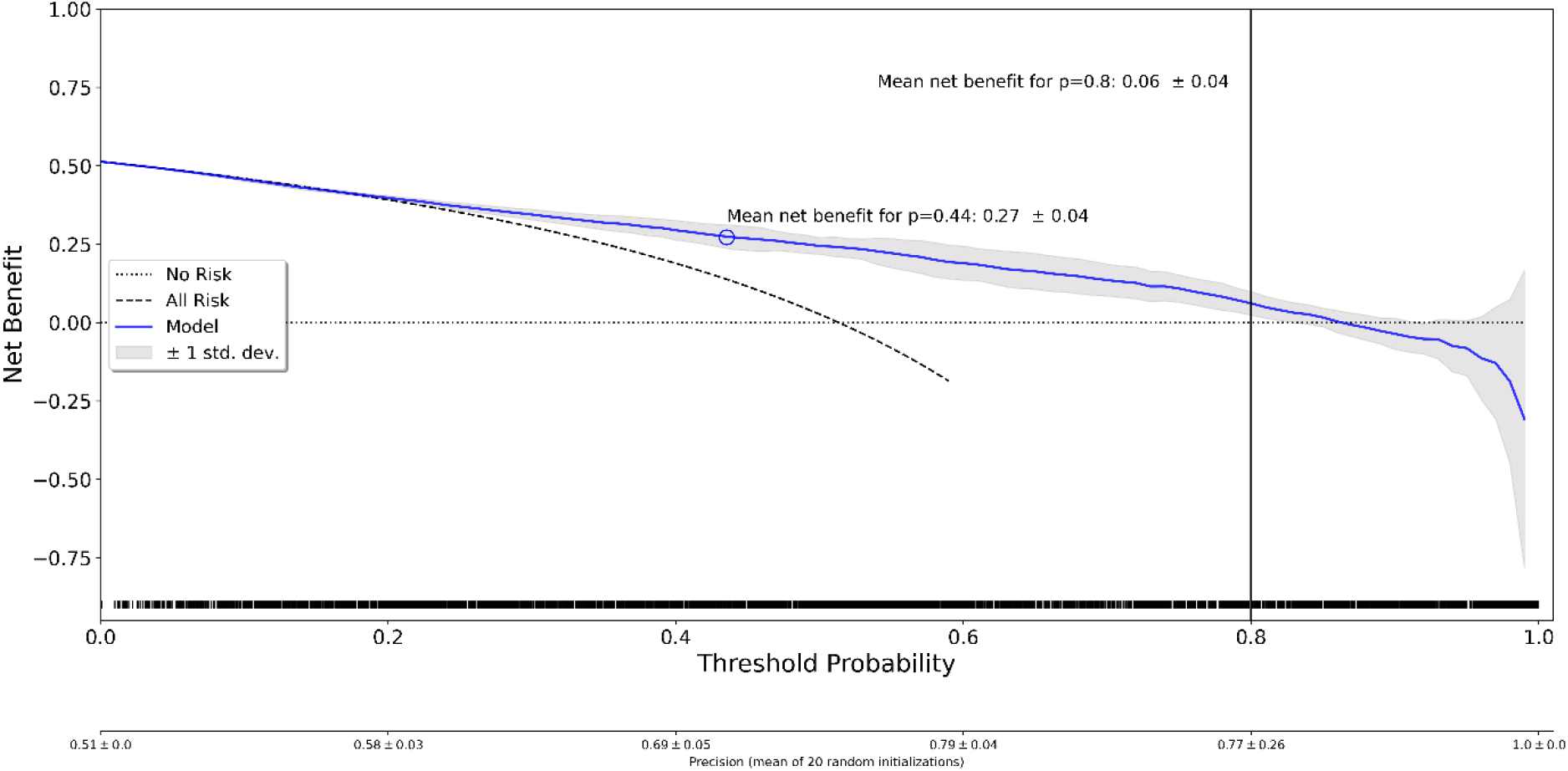
Decision curve analysis for tabular neural network. The blue line indicates the mean and the gray shaded region indicates ± 1 std. band over 20 random initializations. The rug plot shows the samples used for computation of the decision curve. ‘No risk’ denotes the trivial strategy of always predicting mRS 0–2, and ‘All Risk’ denotes the strategy of always predicting mRS 3–6.

## Discussion

While the ability of ML methods to discriminate between good and poor functional outcome looks promising (ROC-AUC ∼0.8), clinical utility in a decision-support scenario aimed at yielding high certainty for prediction of functional dependency (mRS 3–6) is marginal and not evident for the prediction of severe disability or death (mRS 5 and 6). In the latter, observed negative net benefits may even indicate potential harm. Net benefit can be improved when using probability thresholds optimized for F1-scores, suggesting that they may have benefits for other clinical scenarios, in which rating of false-positives and false-negative is more balanced (e.g. information gain for patients or next kin).

In our model, we assessed basic variables available at patient admission, which facilitates the transfer of the developed ML methods to other clinical centers. We verified the result by Nishi et al.^18^ that ML methods can improve prediction of functional outcome when compared to clinical scores. More importantly, we found that most ML methods (with exception of k-NN classifier) provide similar discrimination and calibration performances. Results of primary analysis suggest that the stacked ensemble method performs best with regards to discrimination, calibration and clinical utility. In accordance with previous feature importance analyses reported by Ramos et al.^39^ for ML-based prediction of poor outcome (mRS 5–6) and the analysis by Xu et al.^40^ on predictors of futile recanalization, we found age on admission, NIHSS on admission, and pre stroke independence to be most predictive. In the secondary analysis, all methods showed a considerable drop in performance when used to predict mRS 5-6. This may be attributed to lack of information in baseline variables which differentiate patients with mRS 3–4 from mRS 5–6 patients.

In this study, we aimed at filling the lack of evidence regarding studies on clinical utility of ML models to predict functional outcome in stroke patients. Notably, there is a study available on clinical utility for ML-based outcome prediction in patients with acute ischemic stroke^41^. Jang et al. aimed to compare the clinical utility of ML and logistic regression models for the prediction of functional outcome in a general population of stroke patients. In contrast, the purpose of our study was to investigate the clinical utility of ML models when integrated in an acute stroke workflow to solve a specific task—the prediction of functional dependency and severe disability or death despite successful recanalization (TICI ≥ 2b). The restriction to successfully recanalized patients provided us with a more homogenous patient cohort for model training and provides a potential user with outcome estimates when optimistically assuming that the envisaged intervention is technically successful. Considering a rate of unsuccessful interventions of 15–20%^1,42^, a decision-support algorithm with a cut-off of 80% for predicting poor outcome in successfully reperfusion patients would imply that there is a very high overall chance that the intervention is either not successful or futile (83%–84%, depending on the rate of unsuccessful reperfusion). The 80% cut-off is arbitrary but data on what constitute a sufficiently high certainty for excluding patients from acute stroke reperfusion regimens is not standardized. Certainly, the cut-off needs to be well above 50% (standard cut-off used for reporting on discrimination), to ensure that false positive (classifying patients as potential futile reperfusion, despite they gain functional independence or mRS<5) are weighted more than false negative classifications. Furthermore, our results also show that for more confident cut- offs above 80% the achieved net benefit is further decreasing even resulting in potential harm (negative net benefit). The exact cut-off deemed useful in a clinical scenario may also depend on other factors including health care resources, cultural perception and available information on the patients good will. In summary, the appealing ROC-AUC reported here and by others^39,43^, should thus be handled cautiously with regards to the clinical utility in excluding patients from reperfusion therapies.

We computed a variety of evaluation metrics to provide a detailed description of the ML methods’ performances. In literature, the most frequently reported metric for binary classification problems is the AUC of the ROC curve. However, the ROC-AUC alone can be misleading in case of large class imbalances (i.e. in presence of majority of negative examples). This is also evident in our secondary analysis in which most of the methods achieved similarly good ROC-AUC values as in primary analysis (∼ 0.8) but exhibited a considerably reduced net benefit across all probability thresholds. In addition, the AUC is usually computed over the whole ROC curve taking into account regions which may never be used in practice^44^, whereas the decision curve analysis must be interpreted with respect to clinically relevant thresholds.

The intended use of our ML models is clearly limited to patients selected for treatment with EVT. It is possible that prediction of functional impairment in a more diverse population of patients, which particularly includes individuals not selected for treatment with EVT, may be improved. Nevertheless, the identification of futile recanalization ahead of treatment is of great importance^11^ and we think that our proposed methodology (Figure 2) may serve as a starting point.

In the future, prediction of functional impairment based on patient data available on admission may be improved by incorporation of more elaborate information from laboratory analysis^45,46^ and admission imaging within ML models. The only imaging parameter considered in our feature set was ASPECTS. Several imaging biomarkers have proven to be independently associated with 3-months functional outcome including brain regions affected by the acute lesion^47^, growth of ischemic lesion volume between baseline and 24-hours CT^48^, final infarct volume (based on follow-up CT, 18–36 hours), brain volume^49^, and white matter hyperintensity volume (based on MRI)^50^. In particular, lesion outcome volumes could be predicted ahead of intervention using dedicated deep learning methods^51^ and be incorporated in our proposed ML models.

In conclusion, the clinical utility of ML methods for prediction of functional dependency and severe disability or death despite successful recanalization is marginal when using baseline variables and when considering a clinical decision-support scenario. Further research should be concentrated on the extraction of more elaborate imaging features from admission imaging and incorporation of such in ML prediction models.

## Data Availability

The authors confirm that the data supporting the findings of this study are available within the article and its supplementary materials.

## Footnotes

1 For more details, see https://clinicaltrials.gov/ct2/show/NCT03496064.

