## Supplementary Material for "Beyond Accuracy: Investigating the Potential Clinical Utility of Predicting Functional Dependency and Severe Disability or Death in Successfully Reperfused Patients using Machine Learning"

### Supplementary Materials

**Hyperparameter optimization**

Hyperparameter optimization was performed using a randomized gridsearch (`RandomizedSearchCV`) based on 10-fold stratified cross-validation with 10 iterations (i.e. n_iter=10). Random seeds used to initialize ML algorithms were**:** [42, 84, 126, 168, 210, 252, 294, 336, 378, 420, 462, 504, 546, 588, 630, 672, 714, 756, 798, 840]. All experiments were performed on CPU of laptop computer (Lenovo X1 Extreme with Windows 10, Intel i7-9750H @ 2.6 GHz, 32GB RAM).

*Supplementary Table 1. Hyperparameters of Machine Learning algorithms which were optimized during randomized gridsearch on training data.*

| **Method** | **Hyperparameter** | **Candidate parameter values** | **Optimal parameter mRS 3–6** | **Optimal parameter mRS 5–6** |
|---|---|---|---|---|
| k-NN Classifier | Number of neighbors | [5, 10, 15, 20] | 15 | 5 |
| Linear SVM (liblinear) | C | [0.001, 0.01, 0.1, 1, 10, 100] | 0.1 | 100 |
| | class weight | ['balanced', None] | 'balanced' | None |
| | tolerance | [0.00001, 0.0001, 0.001] | 0.0001 | 0.0001 |
| Linear SVM (libsvm) | C | [0.001, 0.01, 0.1, 1, 10, 100] | 1 | 10 |
| | class weight | ['balanced', None] | None | None |
| | tolerance | [0.00001, 0.0001, 0.001] | 0.0001 | 0.001 |
| Non-linear SVM (libsvm) | C | [0.001, 0.01, 0.1, 1, 10, 100] | 1 | 1 |
| | class weight | ['balanced', None] | None | 'balanced' |
| | tolerance | [0.00001, 0.0001, 0.001] | 0.00001 | 0.0001 |
| Regularized Logistic Regression | C | [0.001, 0.01, 0.1, 1, 10, 100] | 1 | 1 |
| | class weight | ['balanced', None] | None | 'balanced' |
| | tolerance | [0.00001, 0.0001, 0.001] | 0.0001 | 0.0001 |
| | penalty | ['l1', 'l2'] | 'l1' | 'l1' |
| XGB Classifier | Number of estimators | [100, 200, 300, 400, 500] | 200 | 500 |
| | Maximum depth | [3, 4, 5] | 5 | 4 |
| | Minimum number of samples (min_child_weight) | [1, 5, 10] | 1 | 1 |
| | Fraction of samples to subsample | [0.6, 0.8, 1.0] | 0.6 | 0.6 |
| | γ (Minimum loss reduction required to make a further | [0.5, 1, 1.5, 2, 5] | 1.5 | 1 |

| | | | | |
|---|---|---|---|---|
| | partition on a leaf node of the tree) | | | |
| Balanced Random Forest (RF) | Number of estimators | [100, 200, 300, 400, 500] | 500 | 500 |
| | Criterion | ['gini', 'entropy'] | 'gini' | 'gini' |
| | Maximum Features | ['auto', 2, 10] | 'auto' | 2 |
| Multi-layer Perceptron (MLP) | Hidden layer sizes | [(64,), (64, 32), (64, 32, 16)] | (64,) | (64, 32) |
| | Initial learning rate | [0.001, 0.01] | 0.001 | 0.001 |
| | tolerance | [0.00001, 0.0001, 0.001] | 0.001 | 0.001 |
| Stacked ensemble method | | | | |
| Boolean SVM (libsvm) | C | [0.001, 0.01, 0.1, 1, 10, 100] | 0.001 | 1 |
| | class weight | ['balanced', None] | None | 'balanced' |
| | tolerance | [0.00001, 0.0001, 0.001] | 0.00001 | 0.0001 |
| Linear SVM (liblinear) | C | [0.001, 0.01, 0.1, 1, 10, 100] | 0.1 | 10 |
| | class weight | ['balanced', None] | None | 'balanced' |
| | tolerance | [0.00001, 0.0001, 0.001] | 0.00001 | 0.0001 |
| Balanced Random Forest (RF) | Number of estimators | [100, 200, 300, 400, 500] | 400 | 500 |
| | Criterion | ['gini', 'entropy'] | 'gini' | 'entropy' |
| | Maximum Features | ['auto', 2, 10] | 'auto' | 10 |

For tabular neural network, the following hyperparameters were used after empirical evaluation on training data: layers=[64], ps=[0.3], emb_drop=[0.04]. The size of categorical embeddings were computed according to the following heuristics: `emb_szs = {cat: min(600, int(round(1.6 * card**0.56))) for cat, card in zip(categories, cardinalities)}`, with `categories` being a list of categorical variable names and `cardinalities` being the list of respective cardinalities[1]. This resulted in the following network architecture:

```
TabularModel(
  (embeds): ModuleList(
    (0): Embedding(3, 2)
    (1): Embedding(3, 2)
    (2): Embedding(3, 2)
    (3): Embedding(3, 2)
    (4): Embedding(3, 2)
    (5): Embedding(3, 2)
    (6): Embedding(3, 2)
    (7): Embedding(3, 2)
    (8): Embedding(3, 2)
    (9): Embedding(3, 2)
    (10): Embedding(3, 2)
    (11): Embedding(3, 2)
    (12): Embedding(3, 2)
```

[1] Adapted from https://github.com/fastai/fastai1/blob/master/fastai/tabular/data.py#L13

```
    (13): Embedding(5, 3)
    (14): Embedding(4, 3)
    (15): Embedding(4, 3)
    (16): Embedding(4, 3)
    (17): Embedding(3, 2)
    (18): Embedding(3, 2)
  )
  (emb_drop): Dropout(p=0.04, inplace=False)
  (bn_cont): BatchNorm1d(6, eps=1e-05, momentum=0.1, affine=True, track
_running_stats=True)
  (layers): Sequential(
    (0): Linear(in_features=48, out_features=64, bias=True)
    (1): ReLU(inplace=True)
    (2): BatchNorm1d(64, eps=1e-05, momentum=0.1, affine=True, track_ru
nning_stats=True)
    (3): Dropout(p=0.3, inplace=False)
    (4): Linear(in_features=64, out_features=2, bias=True)
  )
)
```

Class-weighted cross entropy with label smoothing was implemented with a smoothing factor ε=0.1 and class weights (0.5, 0.5) for (mRS 0–2, mRS 3–6) and (0.75, 0.25) for (mRS 0–4, mRS 5–6), respectively. Training was performed for 80 epochs using Ranger optimizer with a learning rate of 4e-03 (based on learning rate finder) and flat cosine annealing. Early stopping based on the f1-score in the validation set was employed to prevent overfitting.

**Prediction of Functional Impairment and Severe Disability or Death (primary analysis, mRS 3–6)**

*Supplementary Table 2. Performance for mRS 3–6 prediction. Values are reported as mean ± std. Brier score and ECE are reported for ML algorithms only.*

| Dataset | Method | Accuracy | Balanced Accuracy | Precision | Recall | F1-Score | Specificity | ROC-AUC | AP | Matthews corr. | Brier score | ECE |
|---|---|---|---|---|---|---|---|---|---|---|---|---|
| 10-fold CV | Majority Classifier | 0.52 ± 0.00 | 0.50 ± 0.00 | 0.52 ± 0.00 | 1.0 ± 0.00 | 0.68 ± 0.00 | 0.0 | 0.50 ± 0.00 | 0.52 ± 0.00 | 0.0 | - | - |
| Test data | Majority Classifier | 0.51 | 0.50 | 0.51 | 1.0 | 0.68 | 0.0 | 0.50 | 0.51 | 0.0 | - | - |
| 10-fold CV | SPAN100 Score | 0.64 ± 0.05 | 0.65 ± 0.05 | 0.86 ± 0.10 | 0.36 ± 0.07 | 0.51 ± 0.08 | 0.94 ± 0.04 | 0.75 ± 0.05 | 0.79 ± 0.05 | 0.36 ± 0.11 | - | - |
| Test data | SPAN100 Score | 0.69 | 0.70 | 0.91 | 0.44 | 0.59 | 0.96 | 0.79 | 0.81 | 0.46 | - | - |
| 10-fold CV | PRE Score | 0.65 ± 0.05 | 0.66 ± 0.05 | 0.87 ± 0.09 | 0.38 ± 0.08 | 0.52 ± 0.09 | 0.94 ± 0.03 | 0.77 ± 0.02 | 0.80 ± 0.04 | 0.38 ± 0.11 | - | - |
| Test data | PRE Score | 0.57 | 0.58 | 0.68 | 0.31 | 0.43 | 0.85 | 0.73 | 0.72 | 0.19 | - | - |

| | | | | | | | | | | | | |
|---|---|---|---|---|---|---|---|---|---|---|---|---|
| 10-fold CV | Stroke-TPI Score | 0.71 ± 0.04 | 0.71 ± 0.04 | 0.71 ±0.05 | 0.74 ± 0.04 | 0.72 ± 0.04 | 0.67 ± 0.07 | 0.80 ± 0.03 | 0.83 ± 0.04 | 0.42 ± 0.08 | - | - |
| Test data | Stroke-TPI Score | 0.70 | 0.70 | 0.70 | 0.73 | 0.71 | 0.67 | 0.78 | 0.78 | 0.40 | - | - |
| 10-fold CV | K-NN Classifier | 0.61 ± 0.05 | 0.61 ± 0.05 | 0.65 ± 0.07 | 0.52 ± 0.06 | 0.58 ± 0.05 | 0.69 ± 0.08 | 0.66 ± 0.07 | 0.66 ± 0.07 | 0.22 ± 0.10 | 0.24 ± 0.03 | 0.15 ± 0.03 |
| Test data | K-NN Classifier | 0.66 ± 0.01 | 0.66 ± 0.01 | 0.67 ± 0.01 | 0.66 ± 0.01 | 0.67 ± 0.01 | 0.65 ± 0.01 | 0.72 ± 0.02 | 0.70 ± 0.02 | 0.32 ± 0.01 | 0.22 ± 0.01 | 0.06 ± 0.01 |
| 10-fold CV | Linear SVM (liblinear) | 0.74 ± 0.03 | 0.74 ± 0.03 | 0.76 ± 0.04 | 0.73 ± 0.04 | 0.74 ± 0.03 | 0.75 ± 0.06 | 0.82 ± 0.03 | 0.84 ± 0.04 | 0.48 ± 0.07 | 0.17 ± 0.01 | 0.10 ± 0.02 |
| Test data | Linear SVM (liblinear) | 0.71 ± 0.02 | 0.71 ± 0.02 | 0.69 ± 0.02 | 0.80 ± 0.01 | 0.74 ± 0.02 | 0.63 ± 0.03 | 0.80 ± 0.00 | 0.79 ± 0.01 | 0.43 ± 0.04 | 0.19 ± 0.00 | 0.08 ± 0.01 |
| 10-fold CV | Linear SVM (libsvm) | 0.74 ± 0.03 | 0.74 ± 0.03 | 0.75 ± 0.04 | 0.74 ± 0.05 | 0.74 ± 0.03 | 0.74 ± 0.06 | 0.81 ± 0.03 | 0.83 ± 0.04 | 0.48 ± 0.07 | 0.18 ± 0.01 | 0.11 ± 0.03 |
| Test data | Linear SVM (libsvm) | 0.72 ± 0.02 | 0.72 ± 0.02 | 0.70 ± 0.02 | 0.79 ± 0.02 | 0.74 ± 0.02 | 0.64 ± 0.02 | 0.80 ± 0.01 | 0.79 ± 0.01 | 0.44 ± 0.03 | 0.18 ± 0.00 | 0.08 ± 0.01 |
| 10-fold CV | Non-linear SVM | 0.69 ± 0.04 | 0.69 ± 0.04 | 0.71 ± 0.05 | 0.70 ± 0.07 | 0.70 ± 0.05 | 0.69 ± 0.07 | 0.78 ± 0.03 | 0.80 ± 0.04 | 0.39 ± 0.09 | 0.19 ± 0.01 | 0.11 ± 0.04 |
| Test data | Non-linear SVM | 0.73 ± 0.01 | 0.73 ± 0.01 | 0.71 ± 0.01 | 0.80 ± 0.00 | 0.75 ± 0.01 | 0.65 ± 0.02 | 0.80 ± 0.01 | 0.80 ± 0.01 | 0.48 ± 0.02 | 0.18 ± 0.00 | 0.09 ± 0.00 |
| 10-fold CV | Regularized Logistic Regression | 0.74 ± 0.04 | 0.74 ± 0.04 | 0.75 ± 0.04 | 0.74 ± 0.05 | 0.74 ± 0.04 | 0.74 ± 0.06 | 0.82 ± 0.03 | 0.84 ± 0.04 | 0.47 ± 0.07 | 0.18 ± 0.02 | 0.11 ± 0.02 |
| Test data | Regularized Logistic Regression | 0.70 ± 0.01 | 0.70 ± 0.01 | 0.68 ± 0.01 | 0.79 ± 0.01 | 0.73 ± 0.01 | 0.61 ± 0.02 | 0.79 ± 0.00 | 0.79 ± 0.01 | 0.41 ± 0.03 | 0.19 ± 0.00 | 0.10 ± 0.01 |
| 10-fold CV | XGBoost | 0.73 ± 0.03 | 0.73 ± 0.03 | 0.74 ± 0.04 | 0.72 ± 0.04 | 0.73 ± 0.02 | 0.73 ± 0.06 | 0.81 ± 0.03 | 0.84 ± 0.04 | 0.45 ± 0.05 | 0.18 ± 0.01 | 0.13 ± 0.03 |
| Test data | XGBoost | 0.72 ± 0.01 | 0.72 ± 0.01 | 0.70 ± 0.02 | 0.78 ± 0.02 | 0.74 ± 0.01 | 0.65 ± 0.04 | 0.80 ± 0.01 | 0.80 ± 0.01 | 0.44 ± 0.03 | 0.19 ± 0.01 | 0.09 ± 0.02 |

| | | | | | | | | | | | | |
|---|---|---|---|---|---|---|---|---|---|---|---|---|
| 10-fold CV | Balanced RF | 0.75 ± 0.04 | 0.75 ± 0.04 | 0.77 ± 0.05 | 0.75 ± 0.05 | 0.76 ± 0.04 | 0.76 ± 0.04 | 0.83 ± 0.04 | 0.85 ± 0.05 | 0.51 ± 0.08 | 0.17 ± 0.02 | 0.12 ± 0.02 |
| Test data | Balanced RF | 0.73 ± 0.01 | 0.72 ± 0.01 | 0.70 ± 0.01 | 0.82 ± 0.01 | 0.75 ± 0.01 | 0.63 ± 0.02 | 0.81 ± 0.01 | 0.80 ± 0.01 | 0.46 ± 0.02 | 0.19 ± 0.00 | 0.10 ± 0.01 |
| 10-fold CV | MLP | 0.71 ± 0.04 | 0.71 ± 0.04 | 0.72 ± 0.05 | 0.73 ± 0.05 | 0.72 ± 0.03 | 0.70 ± 0.07 | 0.78± 0.04 | 0.79 ± 0.06 | 0.43 ± 0.07 | 0.21 ± 0.03 | 0.18 ± 0.05 |
| Test data | MLP | 0.73 ± 0.01 | 0.72 ± 0.01 | 0.71 ± 0.02 | 0.78 ± 0.02 | 0.75 ± 0.01 | 0.66 ± 0.03 | 0.80 ± 0.01 | 0.81 ± 0.01 | 0.45 ± 0.02 | 0.19 ± 0.01 | 0.09 ± 0.02 |
| 10-fold CV | Ensemble (Stacking) | 0.74 ± 0.03 | 0.75 ± 0.03 | 0.77 ± 0.04 | 0.73 ± 0.06 | 0.75 ± 0.03 | 0.76 ± 0.05 | 0.83 ± 0.04 | 0.85 ± 0.05 | 0.49 ± 0.06 | 0.17 ± 0.02 | 0.11 ± 0.03 |
| Test data | Ensemble (Stacking) | 0.74 ± 0.01 | 0.74 ± 0.01 | 0.74 ± 0.02 | 0.76 ± 0.02 | 0.75 ± 0.01 | 0.72 ± 0.02 | 0.81 ± 0.00 | 0.81 ± 0.01 | 0.48 ± 0.02 | 0.18 ± 0.01 | 0.06 ± 0.01 |
| 10-fold CV | Tabular Neural Network | 0.78 ± 0.04 | 0.78 ± 0.05 | 0.79 ± 0.05 | 0.78 ± 0.05 | 0.78 ± 0.04 | 0.78 ± 0.07 | 0.82 ± 0.06 | 0.83 ± 0.07 | 0.56 ± 0.09 | 0.17 ± 0.03 | 0.12 ± 0.03 |
| Test data | Tabular Neural Network | 0.73 ± 0.03 | 0.73 ± 0.03 | 0.73 ± 0.03 | 0.75 ± 0.03 | 0.74 ± 0.02 | 0.71 ± 0.05 | 0.80 ± 0.02 | 0.79 ± 0.03 | 0.46 ± 0.05 | 0.18 ± 0.02 | 0.08 ± 0.03 |

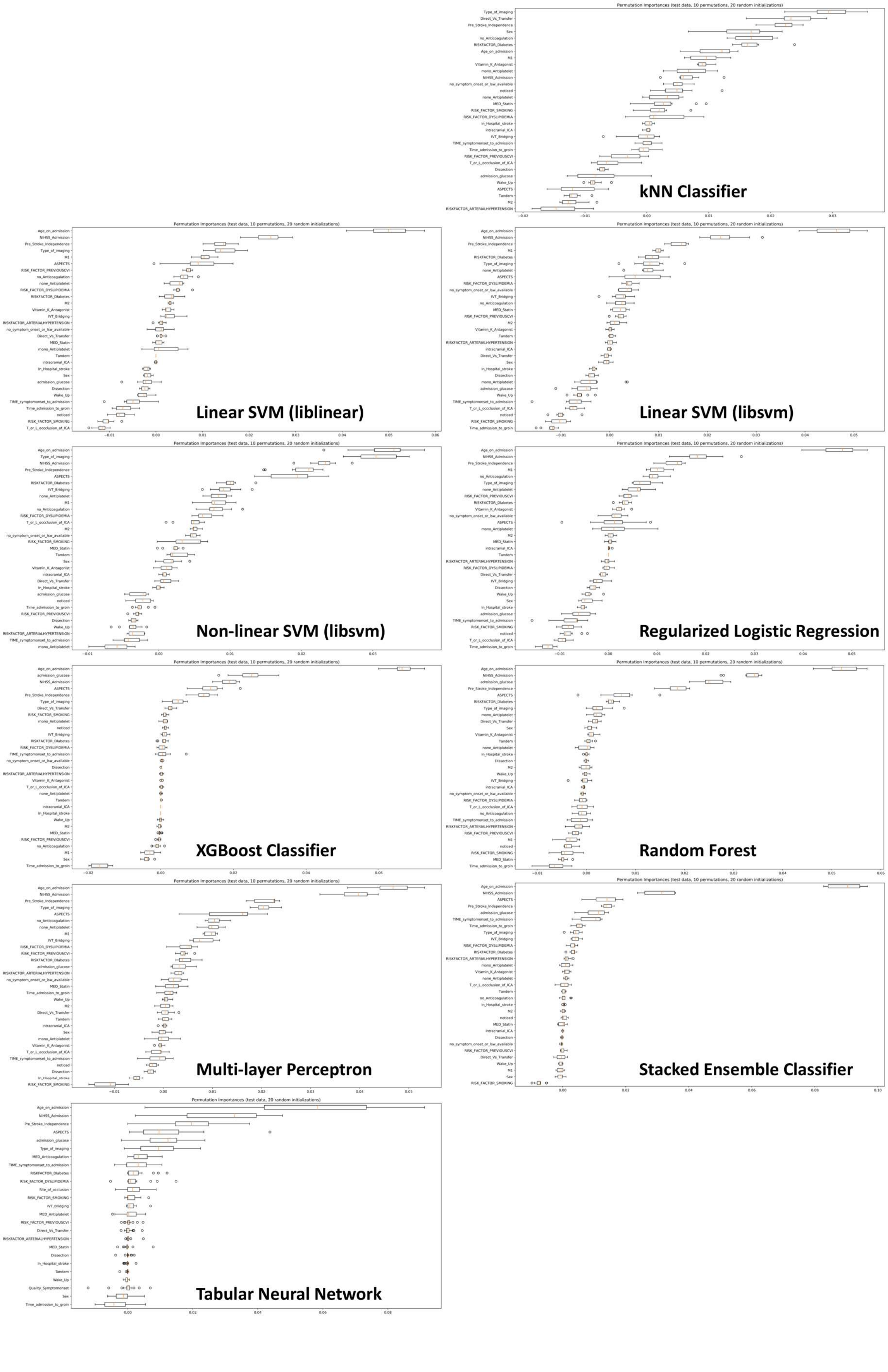


*Supplementary Figure 1. Permutation feature importance computed for all ML methods (mean values of 10 feature permutations and 20 random initializations, primary analysis, mRS 3–6).*

#### Prediction of Functional Impairment and Severe Disability or Death (secondary analysis, mRS 5–6)

*Supplementary Table 3. Performance for mRS 5–6 prediction. Values are reported as mean ± std. Brier score and ECE are reported for ML algorithms only.*

| Dataset | Method | Accuracy | Balanced Accuracy | Precision | Recall | F1-Score | Specificity | ROC-AUC | AP | Matthews corr. | Brier score | ECE |
|---|---|---|---|---|---|---|---|---|---|---|---|---|
| 10-fold CV | Majority Classifier | 0.74 ± 0.01 | 0.50 ± 0.00 | 0.0 | 0.0 | 0.0 | 1.0 ± 0.00 | 0.50 ± 0.00 | 0.26 ± 0.01 | 0.0 | - | - |
| Test data | Majority Classifier | 0.74 | 0.50 | 0.0 | 0.0 | 0.0 | 1.0 | 0.50 | 0.26 | 0.0 | - | - |
| 10-fold CV | SPAN100 Score | 0.77 ± 0.04 | 0.69 ± 0.06 | 0.57 ± 0.08 | 0.51 ± 0.11 | 0.53 ± 0.09 | 0.87 ± 0.03 | 0.73 ± 0.08 | 0.57 ± 0.11 | 0.39 ± 0.11 | - | - |
| Test data | SPAN100 Score | 0.76 | 0.65 | 0.56 | 0.41 | 0.47 | 0.88 | 0.75 | 0.52 | 0.33 | - | - |
| 10-fold CV | PRE Score | 0.76 ± 0.04 | 0.66 ± 0.06 | 0.56 ± 0.10 | 0.45 ± 0.10 | 0.50 ± 0.10 | 0.87 ± 0.03 | 0.73 ± 0.08 | 0.56 ± 0.11 | 0.35 ± 0.13 | - | - |
| Test data | PRE Score | 0.70 | 0.61 | 0.42 | 0.43 | 0.42 | 0.79 | 0.65 | 0.37 | 0.22 | - | - |
| 10-fold CV | Stroke-TPI Score | 0.80 ± 0.04 | 0.66 ± 0.05 | 0.72 ± 0.12 | 0.37 ± 0.09 | 0.49 ± 0.10 | 0.95 ± 0.03 | 0.76 ± 0.07 | 0.62 ± 0.09 | 0.41 ± 0.12 | - | - |
| Test data | Stroke-TPI Score | 0.74 | 0.60 | 0.52 | 0.29 | 0.37 | 0.91 | 0.72 | 0.45 | 0.24 | - | - |
| 10-fold CV | K-NN Classifier | 0.74 ± 0.03 | 0.56 ± 0.06 | 0.48 ± 0.22 | 0.18 ± 0.12 | 0.25 ± 0.14 | 0.94 ± 0.03 | 0.60 ± 0.07 | 0.36 ± 0.06 | 0.17 ± 0.15 | 0.21 ± 0.02 | 0.13 ± 0.04 |
| Test data | K-NN Classifier | 0.76 ± 0.00 | 0.58 ± 0.00 | 0.62 ± 0.00 | 0.20 ± 0.00 | 0.31 ± 0.00 | 0.96 ± 0.00 | 0.66 ± 0.00 | 0.39 ± 0.00 | 0.25 ± 0.00 | 0.19 ± 0.00 | 0.10 ± 0.00 |
| 10-fold CV | Linear SVM (liblinear) | 0.77 ± 0.04 | 0.61 ± 0.06 | 0.63 ± 0.15 | 0.28 ± 0.11 | 0.38 ± 0.13 | 0.94 ± 0.03 | 0.77 ± 0.05 | 0.58 ± 0.08 | 0.30 ± 0.15 | 0.16 ± 0.01 | 0.11 ± 0.02 |
| Test data | Linear SVM (liblinear) | 0.76 ± 0.01 | 0.67 ± 0.01 | 0.55 ± 0.01 | 0.49 ± 0.03 | 0.52 ± 0.02 | 0.86 ± 0.00 | 0.80 ± 0.00 | 0.54 ± 0.00 | 0.36 ± 0.02 | 0.16 ± 0.00 | 0.07 ± 0.01 |
| 10-fold CV | Linear SVM (libsvm) | 0.77 ± 0.04 | 0.62 ± 0.06 | 0.62 ± 0.14 | 0.29 ± 0.11 | 0.39 ± 0.12 | 0.94 ± 0.02 | 0.77 ± 0.06 | 0.59 ± 0.08 | 0.31 ± 0.13 | 0.16 ± 0.01 | 0.11 ± 0.02 |

| | | | | | | | | | | | | |
|---|---|---|---|---|---|---|---|---|---|---|---|---|
| Test data | Linear SVM (libsvm) | 0.77 ± 0.01 | 0.70 ± 0.01 | 0.56 ± 0.02 | 0.54 ± 0.03 | 0.55 ± 0.02 | 0.85 ± 0.01 | 0.80 ± 0.01 | 0.55 ± 0.01 | 0.40 ± 0.02 | 0.16 ± 0.00 | 0.07 ± 0.01 |
| 10-fold CV | Non-linear SVM | 0.76 ± 0.03 | 0.59 ± 0.05 | 0.55 ± 0.22 | 0.23 ± 0.11 | 0.31 ± 0.14 | 0.95 ± 0.02 | 0.74 ± 0.04 | 0.54 ± 0.09 | 0.24 ± 0.15 | 0.16 ± 0.01 | 0.08 ± 0.02 |
| Test data | Non-linear SVM | 0.76 ± 0.01 | 0.63 ± 0.00 | 0.58 ± 0.04 | 0.36 ± 0.01 | 0.45 ± 0.00 | 0.91 ± 0.01 | 0.80 ± 0.01 | 0.56 ± 0.02 | 0.32 ± 0.02 | 0.16 ± 0.00 | 0.05 ± 0.01 |
| 10-fold CV | Regularized Logistic Regression | 0.73 ± 0.04 | 0.73 ± 0.06 | 0.49 ± 0.06 | 0.72 ± 0.11 | 0.58 ± 0.07 | 0.73 ± 0.04 | 0.77 ± 0.06 | 0.59 ± 0.08 | 0.41 ± 0.11 | 0.19 ± 0.02 | 0.20 ± 0.03 |
| Test data | Regularized Logistic Regression | 0.68 ± 0.01 | 0.73 ± 0.01 | 0.44 ± 0.01 | 0.84 ± 0.03 | 0.58 ± 0.01 | 0.63 ± 0.02 | 0.80 ± 0.00 | 0.54 ± 0.00 | 0.41 ± 0.01 | 0.22 ± 0.01 | 0.25 ± 0.01 |
| 10-fold CV | XGBoost | 0.78 ± 0.04 | 0.65 ± 0.06 | 0.62 ± 0.11 | 0.39 ± 0.12 | 0.47 ± 0.12 | 0.92 ± 0.02 | 0.75 ± 0.06 | 0.58 ± 0.09 | 0.36 ± 0.13 | 0.16 ± 0.02 | 0.11 ± 0.03 |
| Test data | XGBoost | 0.75 ± 0.01 | 0.66 ± 0.03 | 0.52 ± 0.03 | 0.46 ± 0.06 | 0.49 ± 0.05 | 0.85 ± 0.01 | 0.80 ± 0.01 | 0.53 ± 0.01 | 0.32 ± 0.05 | 0.16 ± 0.00 | 0.09 ± 0.02 |
| 10-fold CV | Balanced RF | 0.70 ± 0.06 | 0.70 ± 0.08 | 0.46 ± 0.08 | 0.68 ± 0.12 | 0.55 ± 0.09 | 0.71 ± 0.05 | 0.76 ± 0.06 | 0.59 ± 0.09 | 0.36 ± 0.14 | 0.20 ± 0.01 | 0.21 ± 0.02 |
| Test data | Balanced RF | 0.65 ± 0.01 | 0.73 ± 0.01 | 0.42 ± 0.01 | 0.89 ± 0.03 | 0.57 ± 0.01 | 0.57 ± 0.02 | 0.81 ± 0.01 | 0.57 ± 0.03 | 0.40 ± 0.02 | 0.22 ± 0.00 | 0.26 ± 0.01 |
| 10-fold CV | MLP | 0.78 ± 0.03 | 0.64 ± 0.06 | 0.63 ± 0.09 | 0.35 ± 0.12 | 0.44 ± 0.11 | 0.93 ± 0.03 | 0.76 ± 0.05 | 0.57 ± 0.08 | 0.35 ± 0.11 | 0.16 ± 0.02 | 0.11 ± 0.02 |
| Test data | MLP | 0.73 ± 0.03 | 0.68 ± 0.03 | 0.49 ± 0.05 | 0.57 ± 0.09 | 0.52 ± 0.04 | 0.78 ± 0.06 | 0.77 ± 0.02 | 0.50 ± 0.03 | 0.34 ± 0.05 | 0.20 ± 0.03 | 0.15 ± 0.05 |
| 10-fold CV | Ensemble (Stacking) | 0.71 ± 0.06 | 0.70 ± 0.06 | 0.47 ± 0.09 | 0.68 ± 0.08 | 0.55 ± 0.07 | 0.72 ± 0.07 | 0.78 ± 0.07 | 0.61 ± 0.09 | 0.37 ± 0.11 | 0.19 ± 0.02 | 0.20 ± 0.03 |
| Test data | Ensemble (Stacking) | 0.71 ± 0.02 | 0.72 ± 0.02 | 0.46 ± 0.02 | 0.74 ± 0.04 | 0.57 ± 0.02 | 0.70 ± 0.02 | 0.79 ± 0.01 | 0.54 ± 0.02 | 0.39 ± 0.03 | 0.19 ± 0.01 | 0.19 ± 0.01 |
| 10-fold CV | Tabular Neural Network | 0.76 ± 0.05 | 0.75 ± 0.05 | 0.53 ± 0.07 | 0.74 ± 0.08 | 0.62 ± 0.07 | 0.76 ± 0.06 | 0.78 ± 0.07 | 0.61 ± 0.09 | 0.46 ± 0.10 | 0.19 ± 0.02 | 0.22 ± 0.02 |

| | | | | | | | | | | | | |
|---|---|---|---|---|---|---|---|---|---|---|---|---|
| Test data | Tabular Neural Network | 0.74 ± 0.03 | 0.71 ± 0.04 | 0.51 ± 0.05 | 0.65 ± 0.06 | 0.57 ± 0.05 | 0.77 ± 0.03 | 0.78 ± 0.04 | 0.56 ± 0.05 | 0.40 ± 0.07 | 0.18 ± 0.01 | 0.16 ± 0.02 |

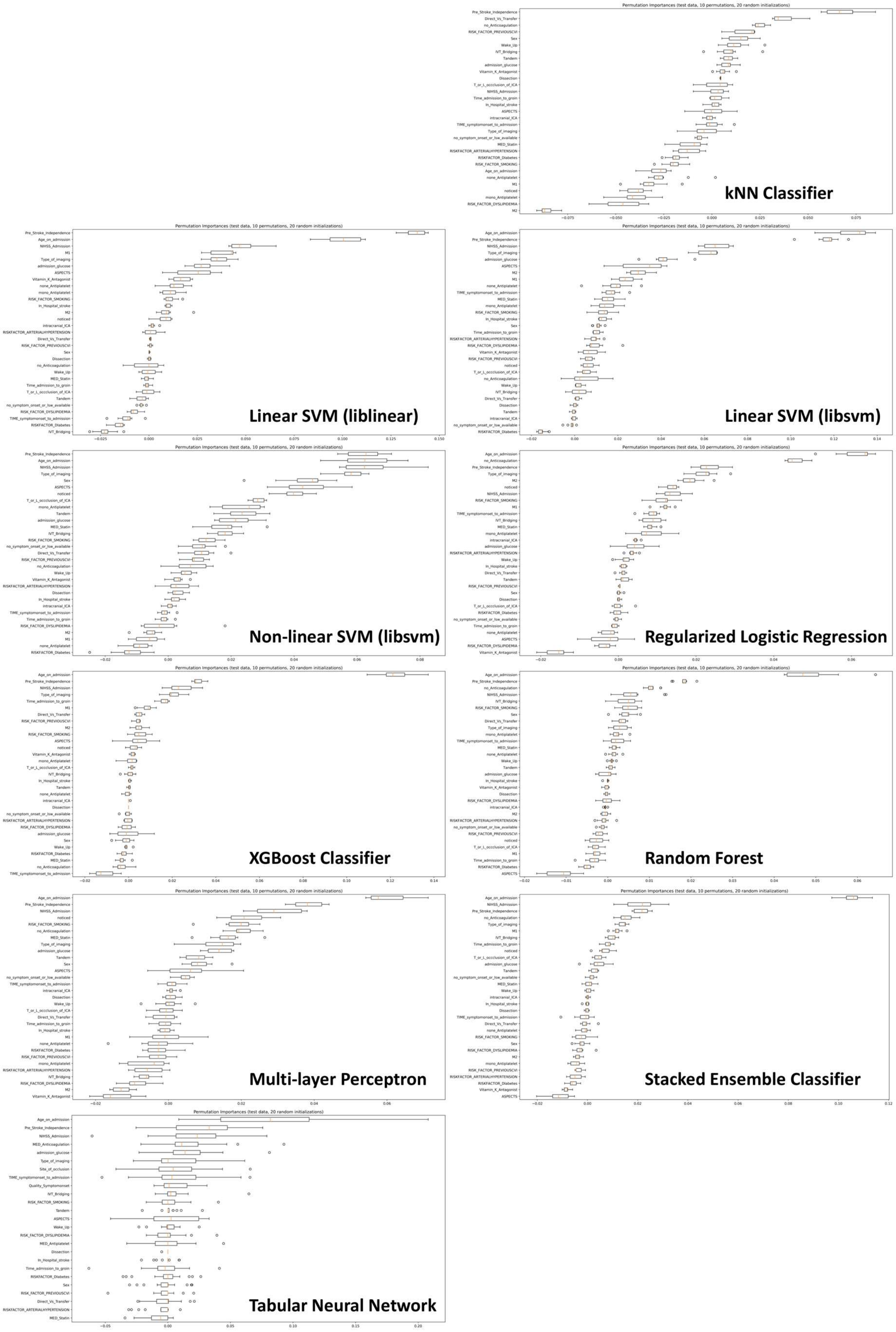


*Supplementary Figure 2. Permutation feature importance computed for all ML methods (mean values of 10 feature permutations and 20 random initializations, secondary analysis, mRS 5–6).*

#### Clinical Utility of Machine Learning Models (primary analysis, mRS 3–6)

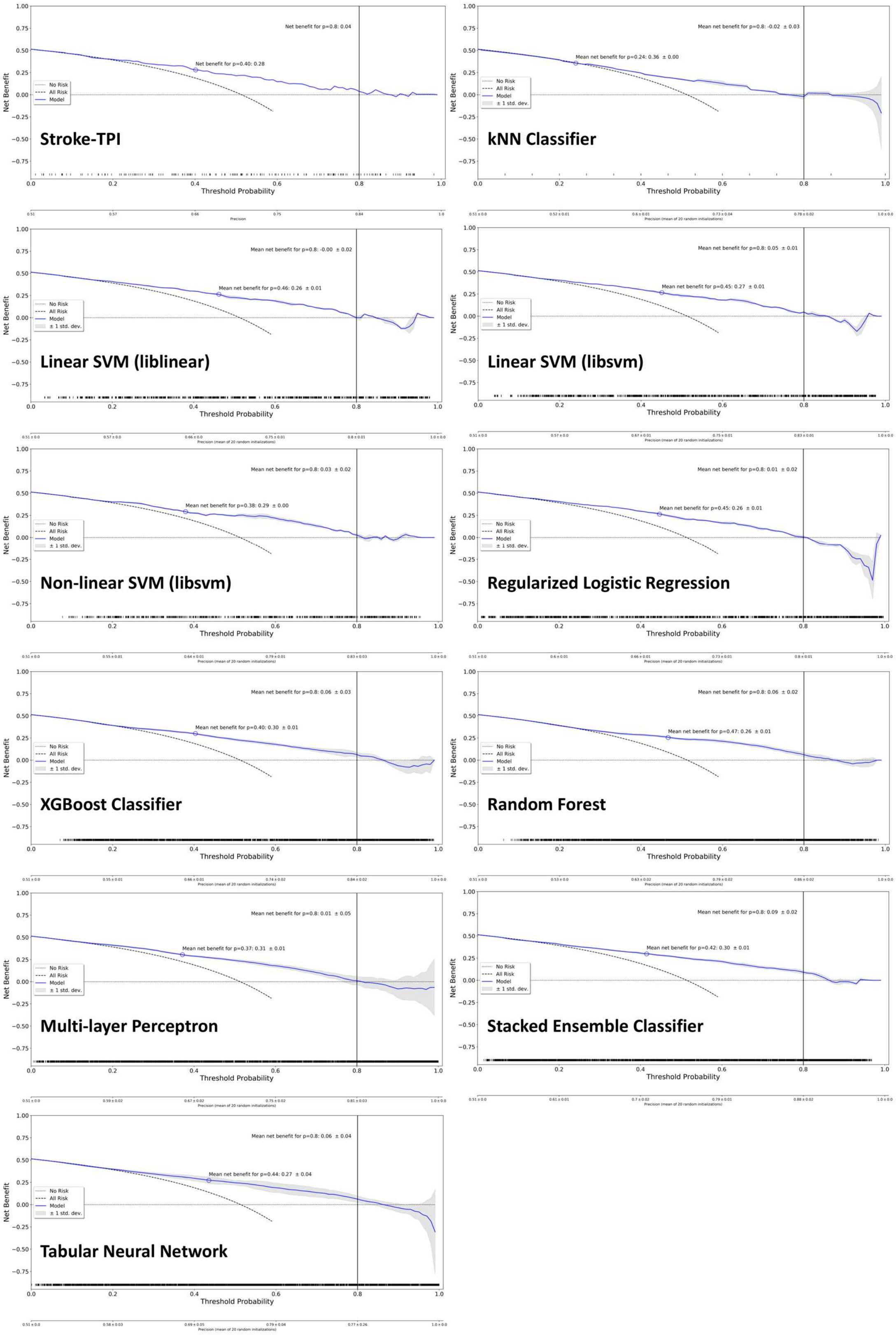


*Supplementary Figure 3. Decision curve analyses for Stroke-TPI and all ML methods (primary analysis, mRS 3–6).*

**Clinical Utility of Machine Learning Models (secondary analysis, mRS 5–6)**

*Supplementary Table 4. Results of decision curve analyses for different ML algorithms (and Stroke-TPI score) in case of mRS 5–6 prediction.*

| Method | Mean Net Benefit for F1-optimized threshold | Mean Net Benefit for p=0.8 |
|---|---|---|
| Stroke-TPI score | 0.05 ± 0.00 for p=0.35 | -0.02 ± 0.00 |
| k-NN Classifier | 0.08 ± 0.00 for p=0.22 | -0.04 ± 0.00 |
| Linear SVM (liblinear) | 0.09 ± 0.01 for p=0.34 | -0.01 ± 0.01 |
| Linear SVM (libsvm) | 0.10 ± 0.01 for p=0.32 | -0.01 ± 0.00 |
| Non-linear SVM | 0.08 ± 0.01 for p=0.32 | 0.00 ± 0.00 |
| Regularized Logistic Regression | -0.09 ± 0.02 for p=0.57 | -0.18 ± 0.06 |
| XGBoost | 0.11 ± 0.01 for p=0.29 | -0.08 ± 0.02 |
| Balanced Random Forest | -0.06 ± 0.03 for p=0.56 | -0.05 ± 0.06 |
| MLP | 0.06 ± 0.01 for p=0.35 | -0.23 ± 0.14 |
| Ensemble (Stacking) | -0.03 ± 0.02 for p=0.57 | -0.04 ± 0.02 |
| Tabular Neural Network | 0.00 ± 0.02 for p=0.54 | -0.06 ± 0.04 |

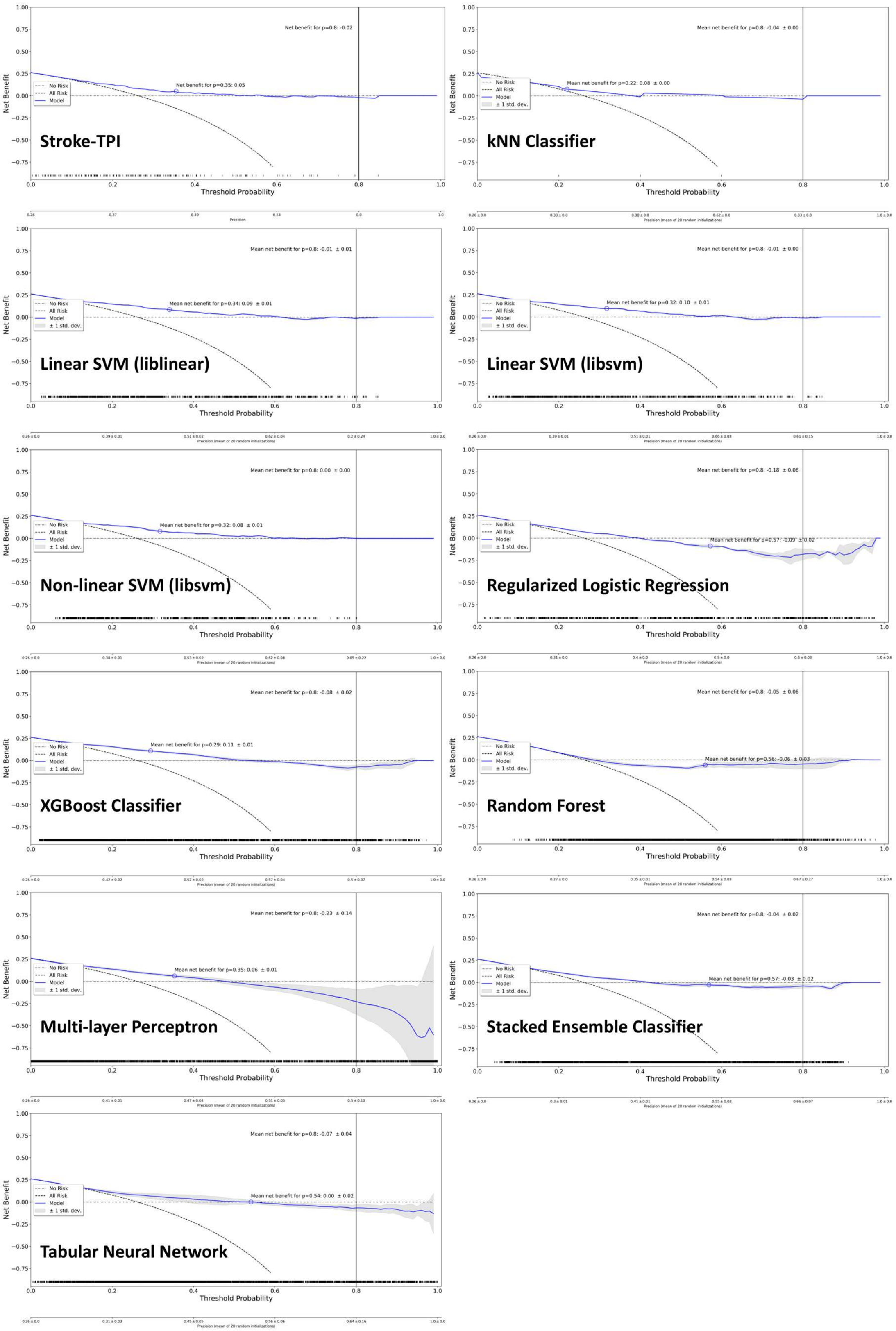


*Supplementary Figure 4. Decision curve analyses for Stroke-TPI and all ML methods (secondary analysis, mRS5–6).*
